# Emergence of the SARS-CoV-2 B.1.1.7 lineage and its characteristics at an outpatient testing site in Berlin, Germany, January-March 2021

**DOI:** 10.1101/2021.04.15.21255389

**Authors:** Welmoed van Loon, Heike Rössig, Susen Burock, Jörg Hofmann, Julian Bernhardt, Elizabeth Linzbach, Domenika Pettenkofer, Christian Schönfeld, Maximilian Gertler, Joachim Seybold, Tobias Kurth, Frank P. Mockenhaupt

## Abstract

Within five weeks in early 2021, B.1.1.7 became the dominant SARS-CoV-2 lineage at an outpatient testing site in Berlin. Characteristics including Ct-values of 193 and 125 recently ill outpatients with B.1.1.7 and wildtype virus, respectively, were similar, except for more commonly reported sore throat and travel, and less frequently stated loss of smell and taste in the former.

---

The SARS-CoV-2 B.1.1.7 lineage (variant of concern [VOC] 202012/01, or 20I/501Y.V1) likely emerged during autumn 2020 in the United Kingdom and quickly became the dominant strain (1). It carries multiple mutations and deletions, including 501Y and deletion ΔH69/ΔV70 (del69–70) in the spike protein. The B.1.1.7 variant is considered to exhibit increased transmissibility compared to non-VOC lineages (1), hereafter referred to as wildtype virus, whereas increased fatality is ambiguous (2,3).

At the outpatient SARS-CoV-2 testing site of Charité - Universitätsmedizin Berlin, the first patient infected with the B.1.1.7 variant was identified on January 18, 2021. Herein, we describe lineage prevalence over time as well as demographic and clinical characteristics in outpatients with the B.1.1.7 variant and those with the wildtype virus presenting until end of March. Ethical approval for the analysis was obtained from Charité’s institutional review board (EA4/083/20).

## The study

Details of the testing site have been described (4). Upon presentation, physicians interviewed patients on demographics, medical history, and symptoms. If indicated, an combined oro-nasopharyngeal swab was collected. SARS-CoV-2 infection was assessed applying a cobas® 6800/8800 analyzer (Roche Diagnostics, Mannheim, Germany), targeting both *ORF1ab* and *E Gene* (5). All positive samples were typed for the N501Y and del69–70 polymorphisms by melting curve analysis. Variants including both polymorphisms were considered B.1.1.7.

Between January 18 and March 29, 2021, 349 SARS-CoV-2 positive patients presented. In this period, the proportion of the B.1.1.7 variant increased from 2% to >90% (Figure 1). In total, 35.8% (125/349) of the samples belonged to the wildtype lineage, 57.0% (199/349) were the B.1.1.7 variant, and 7.2% (25/349) other non-wildtype variants (non-VOCs).

**Figure 1.**
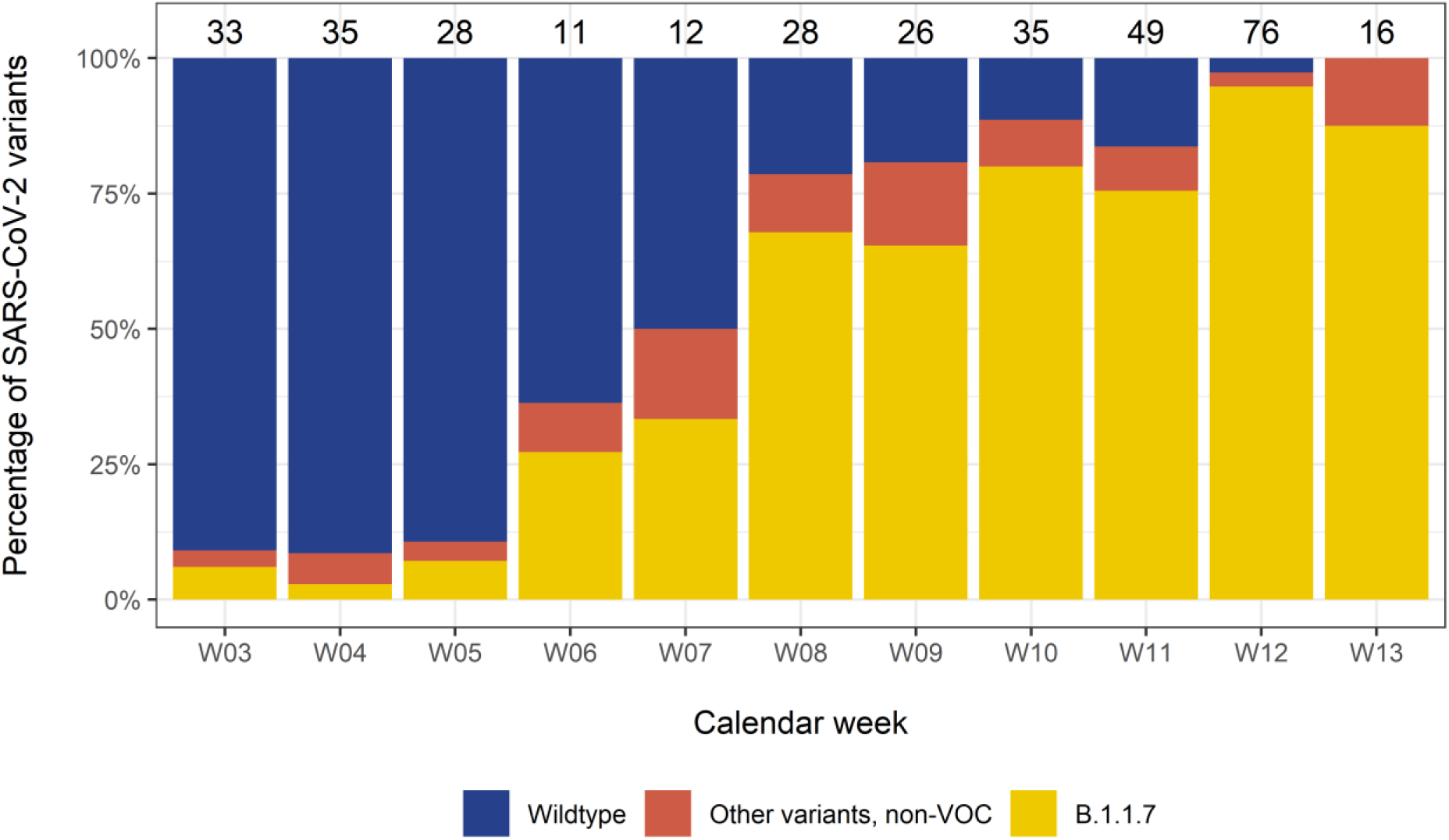
Proportion of SARS-CoV-2 lineages at the Charité testing site, 2021. The numbers on top of the bars indicate the total number of positive SARS-CoV-2-tests assessed. Six (partially) vaccinated outpatients are included for completeness. Note that calendar week 13 only includes one day (March 29).

Six patients previously had received ≥1 SARS-CoV-2 vaccination (five, first dose AZD1222 [Oxford-AstraZeneca] 4-24 days before; one, second BNT162b2 [Pfizer/BioNTech] dose 66 days before); all were infected with B.1.1.7 and were excluded from analysis as were patients carrying lineages other than wildtype or B.1.1.7. Half of the patients were female (49%); mean age was 36±15 years. Almost all reported symptoms (97%). Median symptom duration until testing was 3 days (IQR, 2, 4). Leading complaints were fatigue (72%), headache (69%), and muscle ache (60%). Fifteen percent reported travel outside of Berlin in the last fourteen days, and half (49%) had had contact with a confirmed SARS-CoV-2 case (Table 1).

**Table 1.** Characteristics of SARS-CoV-2 positive outpatients attending the Charité testing site, separated by wildtype and B.1.1.7 lineages.

|  | Wildtype lineage<br>N=125<br>(%, mean [SD], or<br>median [25<sup>th</sup>, 75<sup>th</sup><br>quantile]) | B.1.1.7 variant<br>N=193<br>(%, mean [SD], or<br>median [25<sup>th</sup>, 75<sup>th</sup><br>quantile]) | OR [95% CI],<br>difference in mean<br>[95% CI], or<br>difference in<br>median [95% CI]* |
| --- | --- | --- | --- |
| Female | 44.8% | 51.0% | 1.3 (0.8, 2.0) |
| Age, years | 36.6 (13.8) | 34.8 (15.9) | 1.8 (-1.5, 5.1) |
| Any symptoms | 97.6% | 96.4% | 0.7 (0.2, 2.6) |
| Self-reported fever in the<br>last 48 hours | 38.4% | 42.5% | 1.2 (0.8, 1.9) |
| Temperature in case of<br>fever (°C, by patient<br>statement) | 38.3 (0.6) | 38.2 (0.7) | 0.1 (-0.2, 0.4) |
| Short of breath | 9.6% | 13.5% | 1.5 (0.7, 3.0) |
| Fatigue | 73.6% | 71.5% | 0.9 (0.5, 1.5) |
| Pain on the chest | 2.4% | 1.0% | 0.4 (0.1, 2.6) |
| Diarrhea | 15.2% | 12.4% | 0.8 (0.4, 1.5) |
| Loss of smell/taste | 37.6% | 23.8% | 0.5 (0.3, 0.9) |
| Muscle ache | 60.0% | 60.1% | 1.0 (0.6, 1.6) |
| Sore throat | 41.6% | 53.9% | 1.6 (1.0, 2.6) |
| Cough | 48.8% | 50.8% | 1.1 (0.7, 1.7) |
| Headache | 68.8% | 68.9% | 1.0 (0.6, 1.6) |
| Chills | 35.2% | 34.7% | 1.0 (0.6, 1.6) |
| Rhinorrhea | 60.8% | 52.8% | 0.7 (0.5, 1.1) |
| Symptom duration upon<br>test (days) | 3.0 (2.0, 4.8) | 3.0 (2.0, 4.0) | 0.0 (-1.0, 0.0) |
| Contact to a confirmed<br>SARS-CoV-2 case | 48.0% | 50.3% | 1.1 (0.7, 1.7) |
| Time between contact to a<br>confirmed SARS-CoV-2<br>case and test (days) | 4.0 (1.2, 7.0) | 4.0 (1.0, 6.0) | 0.0 (-1.5, 3.0) |
| Traveled outside the | 9.6% | 18.7% | 2.2 (1.1, 4.3) |

|  | Wildtype lineage | B.1.1.7 variant | OR [95%CI],<br>difference in mean<br>[95%CI], or<br>difference in<br>median [95%CI]* |
| --- | --- | --- | --- |
|  | N=125 | N=193 |  |
|  | (%, mean [SD], or<br>median [25<sup>th</sup>, 75<sup>th</sup><br>quantile]) | (%, mean [SD], or<br>median [25<sup>th</sup>, 75<sup>th</sup><br>quantile]) |  |
| Berlin region in the last<br>14 days |  |  |  |
| Ct-value | 20.2 (17.4, 24.1) | 20.1 (17.1, 22.8) | 0.1 (-0.9, 1.6) |
| Symptom duration < 7<br>days | N=113;<br>19.9 (17.4, 23.5) | N =171;<br>19.5 (16.6, 22.6) | 0.4 (-1.0, 1.7) |
| Symptom duration ≥<br>7 days | N=6;<br>30.1 (26.1, 31.3) | N=11;<br>26.2 (21.4, 31.2) | 3.9 (-5.6, 10.0) |

\* The 95% confidence interval for the difference in medians was computed by a percentile bootstrap with 1,000 replications.

Most assessed characteristics did not substantially differ between patients carrying the SARS-CoV-2 wildtype or the B.1.1.7 lineage, including age, sex, leading complaints, symptom duration, contacts to a confirmed case, and time passed since such contacts (Table 1). However, B.1.1.7 patients had travelled more frequently than those infected with the wildtype strain (19% *vs*. 10%). As for symptoms, patients with B.1.1.7 more often reported sore throat than those with the wildtype lineage (54% *vs*. 42%) but less often loss of smell/taste (24% *vs*. 38%) (Table 1).

We observed no difference in cycle threshold (Ct)-values for the *E Gene* target between B.1.1.7 and wildtype samples (medians, 20.2 *vs*. 20.1) (Table 1). In patients reporting a symptom duration of ≥7 days, Ct-values appeared to be lower for the B.1.1.7 lineage (median, 26.2; wildtype, 30.1; Mann-Whitney-U-test, *P=*0.7) (Table 1, Figure 2).

**Figure 2.**
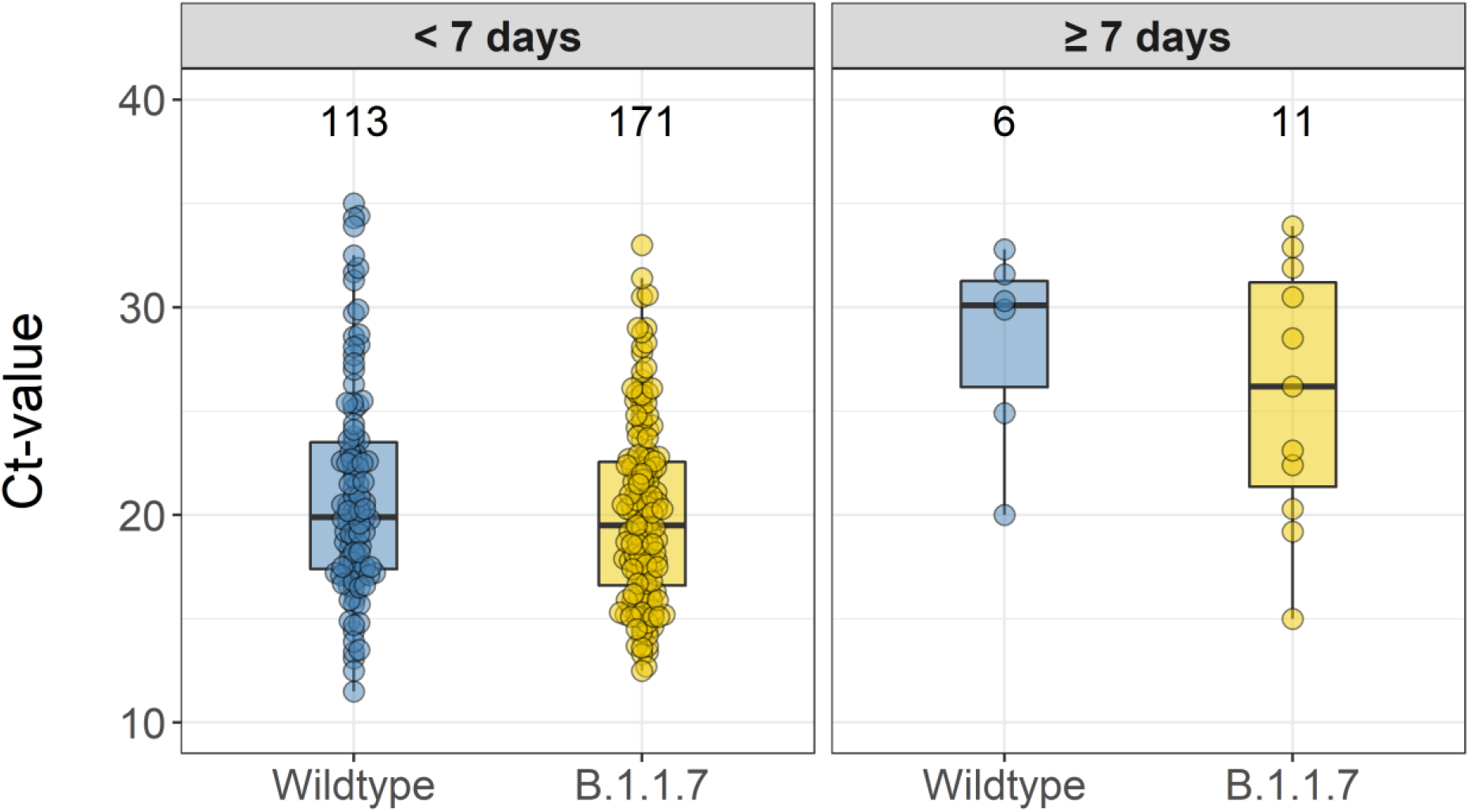
Comparison of median Ct-values in SARS-CoV-2 wildtype and B.1.1.7 lineage, by symptom duration. The boxplots indicate medians, 25^th^ and 75^th^ percentiles (i.e., Q1 and Q3). The upper whiskers reach the largest value with a maximum Q3 + 1.5 * interquartile range (IQR). The lower whiskers reach the smallest value with a minimum Q1 - 1.5 * IQR. The numbers on top of the boxplots indicate the total number of observations included in the comparison.

Lastly, we explored which combination of variables in our dataset best predicted the B.1.1.7 variant in a logistic regression applying a backward stepwise selection based on the Akaike information criterion. This identified the best set of predictors as absent loss of smell/taste (*P=0*.*01*), longer symptom duration (*P=0*.*02*), sore throat (*P=0*.*05*), lower Ct-value (*P=0*.*07*), travel in the last 14 days (*P=0*.*08*), lower age (*P=0*.*09*), and absent rhinorrhea (*P=0*.*12*). We then used bootstrap technique to repeat the variable selection in 1,000 replicated datasets and evaluated how often these variables were selected with the backward selection. This resulted in loss of smell/taste, 89%; symptom duration, 78%; travel, 73%; sore throat, 68%; Ct-value, 66%; age, 56%; rhinorrhea, 51%. Absent loss of smell/taste, longer symptom duration, and travel were selected most often, indicating their importance in B.1.1.7 variant prediction. All analyses were done in R version 3.6.3.

## Discussion

The first B.1.1.7 case in Germany was notified in late December 2020 (6). At our testing site, the B.1.1.7 lineage was first observed three weeks later and replaced the wildtype virus as the dominant strain within five weeks only. The rapid emergence and dominance of this lineage likely results from its increased transmissibility (1), which is potentially caused by spike protein polymorphisms including 501Y conferring enhanced mucosal binding (7); 681H, near a region important for transmission (8); deletion 69-70, linked to immune escape (9). Viral replication *in vitro* does not differ from pre-existing strains (10).

The B.1.1.7 variant reportedly is associated with increased mortality (2), although this has been questioned (3). In our young outpatient study population, we did not observe major, lineage-dependent differences in leading symptoms. Nevertheless, loss of smell/taste, among the most specific COVID-19 symptoms (4), was less common in patients infected with B.1.1.7., whereas, sore throat was more frequently reported than in wildtype virus patients. Similarly, a UK survey revealed less loss of smell/taste in B.1.1.7 patients but more sore throat, cough, fatigue, myalgia, and fever (11). In contrast, no associations between SARS-CoV-2 B.1.1.7 and self-reported symptoms, disease duration, or hospital admissions were seen in another UK study (12). The most important factors for B.1.1.7 infection prediction in our study appeared to be loss of smell/taste and longer symptom duration, in addition to recent travel.

As for Ct-values, one study observed similar figures in patients infected with B.1.1.7 and wildtype lineages; however, a longer duration of infection with the B.1.1.7 variant was suggestive by repeated sampling over time (13). Likewise, longer persistence has been observed for B.1.1.7 (14), but also lower Ct-values (higher viral load) compared to wildtype samples. Lower Ct-values were also seen in other studies on population (15) and inpatient levels (3). In our cross-sectional assessment of recently ill outpatients, we did not observe such differences. Still, increased transmissibility may result from the variant’s prolonged excretion (13,14). Test timing appears crucial for the interpretation of Ct-values. Outpatients are commonly tested earlier in the course of infection than inpatients. The combination of prolonged viral shedding with different test timing might explain increased viral load in B.1.1.7 samples in inpatients, but not in recently ill outpatients. In support of that, in our outpatients with ≥7 days symptom duration, Ct-values in B.1.1.7 samples were suggestively reduced. However, the comparison groups were small. Our data enable to detect a maximum effect size on overall Ct-values, which corresponds to B.1.1.7 Ct-values being 1.5 units below wildtype, or 1.0 above.

The main limitation in our study are limited subgroup sizes, which reduced the likelihood to detect differences between rare characteristics. Others are the one-time assessment, subjective symptom duration, and the variable manifestation of SARS-CoV-2 infection (4). Strengths include standardized procedures conducted by trained medical staff and the prospective nature of the study evaluating patient groups during the same time period, reducing the likelihood of confounding due to temporal effects.

SARS-CoV-2 VOC B.1.1.7 is now the dominant lineage in Berlin. On the outpatient level, there appears to be no major difference in clinical manifestation.

## Data Availability

The data presented in this study are available on reasonable request from the corresponding author.

## Acknowledgments

We thank the Corona Untersuchungsstelle (CUS) staff at the Charité - Universitaetsmedizin Berlin for their dedication and hard work during the COVID-19 pandemic.

